# Fronto-Striatal Dynamic Connectivity is linked to Dopaminergic Motor Response in Parkinson’s Disease

**DOI:** 10.1101/2022.09.29.22280487

**Authors:** Lukas Hensel, Aline Seger, Ezequiel Farrher, Anna K. Bonkhoff, N. Jon Shah, Gereon R. Fink, Christian Grefkes, Michael Sommerauer, Christopher E. J. Doppler

## Abstract

**Background:** Differences in dopaminergic motor response in Parkinson’s disease (PD) patients are related to specific PD subtypes. An important factor driving dopaminergic response might lie in the temporal dynamics in corticostriatal connections.

**Objectives:** The aim of this study is to determine if altered resting-state dynamic functional network connectivity (dFNC) is associated with dopaminergic motor response.

**Methods:** We assessed static and dFNC in 32 PD patients and 18 healthy controls (HC). Patients were subgrouped according to their dopaminergic motor response as low and high responders using a median split.

**Results:** Patients featuring high dopaminergic responses spent more time in a regionally more integrated state 1 compared to HC. Furthermore, dFNC between aMCC/dACC (anterior midcingulate cortex/dorsal anterior cingulate cortex) and putamen was lower in low responders during a more segregated state 2 and correlated with dopaminergic motor response.

**Conclusions:** Alterations in temporal dynamics of fronto-striatal connectivity might underlie treatment response in PD.

## Introduction

Parkinson’s disease (PD) is characterized by a striatal dopaminergic depletion due to neurodegeneration, yet often a considerable response of motor symptoms to dopaminergic replacement therapy can be observed. Even though this hallmark is part of the diagnostic criteria of PD, a substantial number of patients only show a limited response to dopaminergic therapy in clinical practice (1,2), even in autopsy-proven PD cases (1,2). The underlying mechanism for these differences is poorly understood. Using functional magnetic resonance imaging (fMRI), dopaminergic medication was shown to impact on connectivity of both cortico-cortical and cortico-striatal connections, indicating that dopaminergic response lies in interindividual differences in motor network connectivity (3). Previous resting state fMRI (rs-fMRI) investigations have suggested that higher functional connectivity of the striatum with remote cortical motor centers facilitates dopaminergic response (4–6). Importantly, the majority of studies on connectivity changes in PD have measured the mean connectivity over a longer duration, i.e., static connectivity (4). However, novel developments in analysing fMRI connectivity data allow to assess transient changes of connectivity, providing a more detailed picture of the brain’s physiology in health and disease (7). For example, recent studies using a dynamic functional network connectivity (dFNC) approach were able to link changes of dFNC with cognitive impairment and higher non-motor burden (8,9), suggesting distinct connectivity patterns according to PD subtypes (10). Yet, it remains unclear how alterations of dFNC relate to the response to dopaminergic therapy. Therefore, we investigated if dFNC could distinguish between PD patients with low and high treatment response.

## Materials and methods

### Participants

Thirty-two PD patients, diagnosed according to the Movement Disorder Society (MDS) criteria (11), and 18 age- and sex-matched controls underwent rs-fMRI. Inclusion criteria were age between 51 - 80 years, Geriatric Depression Scale (GDS-15) < 5, and Montreal Cognitive Assessment (MoCA) score > 22. Exclusion criteria encompassed contraindications for MRI, and - for patients - a motor symptom duration > 15 years. Motor deficits were assessed using the MDS Unified Parkinson’s Disease Rating Scale part III (MDS-UPDRS III) during patients’ regular ON state and during OFF state after 12 hours without dopaminergic medication. To determine the dopaminergic treatment response, the ratio between ON and OFF states was calculated. Patients were subgrouped using a median split of the response to dopaminergic therapy, distinguishing low and high responders (median = 26%). Additionally, we recorded motor symptom duration, Hoehn and Yahr stage, and levodopa equivalent daily doses (LEDD) (12).

Details about the neuropsychological assessment are given in the supplementary material. The study was approved by the local ethics committee and all study participants gave written informed consent according to the Declaration of Helsinki.

### Magnetic resonance imaging acquisition

Magnetic resonance images were acquired using a Siemens Trio 3T MR scanner. For rs-fMRI, participants were instructed to close their eyes and let their minds wander. Patients were instructed not to fall asleep, which was verified upon the end of the scan. An echo planar imaging (EPI) sequence comprising 300 volumes was utilized with the following parameters: repetition-time (TR) = 2200 ms, echo-time (TE) = 30 ms, field-of-view (FoV) = 200×200 mm^2^, 36 axial slices, 3.1 mm^3^ isotropic voxel-size, flip-angle = 90°. Additionally, T1-images were acquired (MP-RAGE, TR = 2.5 s, TE = 2.89 ms, FoV = 256×232 mm^2^, 176 axial slices, 1 mm^3^ isotropic voxel-size, flip-angle = 7°) to screen for structural abnormalities.

### Static and Dynamic Connectivity

Details about data preprocessing are given in the supplementary material. Functional connectivity was extracted using the GIFT toolbox (version 4.0, https://trendscenter.org/software/gift/). First, we used an independent component analysis (ICA), constrained by components from rs-fMRI data of 405 healthy controls (13,14), to extract spatially distinct intrinsic components (15,16). From these components, ten volumes of interest (VOIs) were selected, contributing to four key motor systems (17), namely the sensorimotor network, the motor initiation network, the basal ganglia network and the cerebellar network (Figure 1 A). For artifact removal, components’ time courses were detrended, despiked using 3Ddespike (18), filtered by a fifth-order Butterworth low-pass filter (cut-off: 0.15 Hz), and normalized (19).

**Figure 1:**
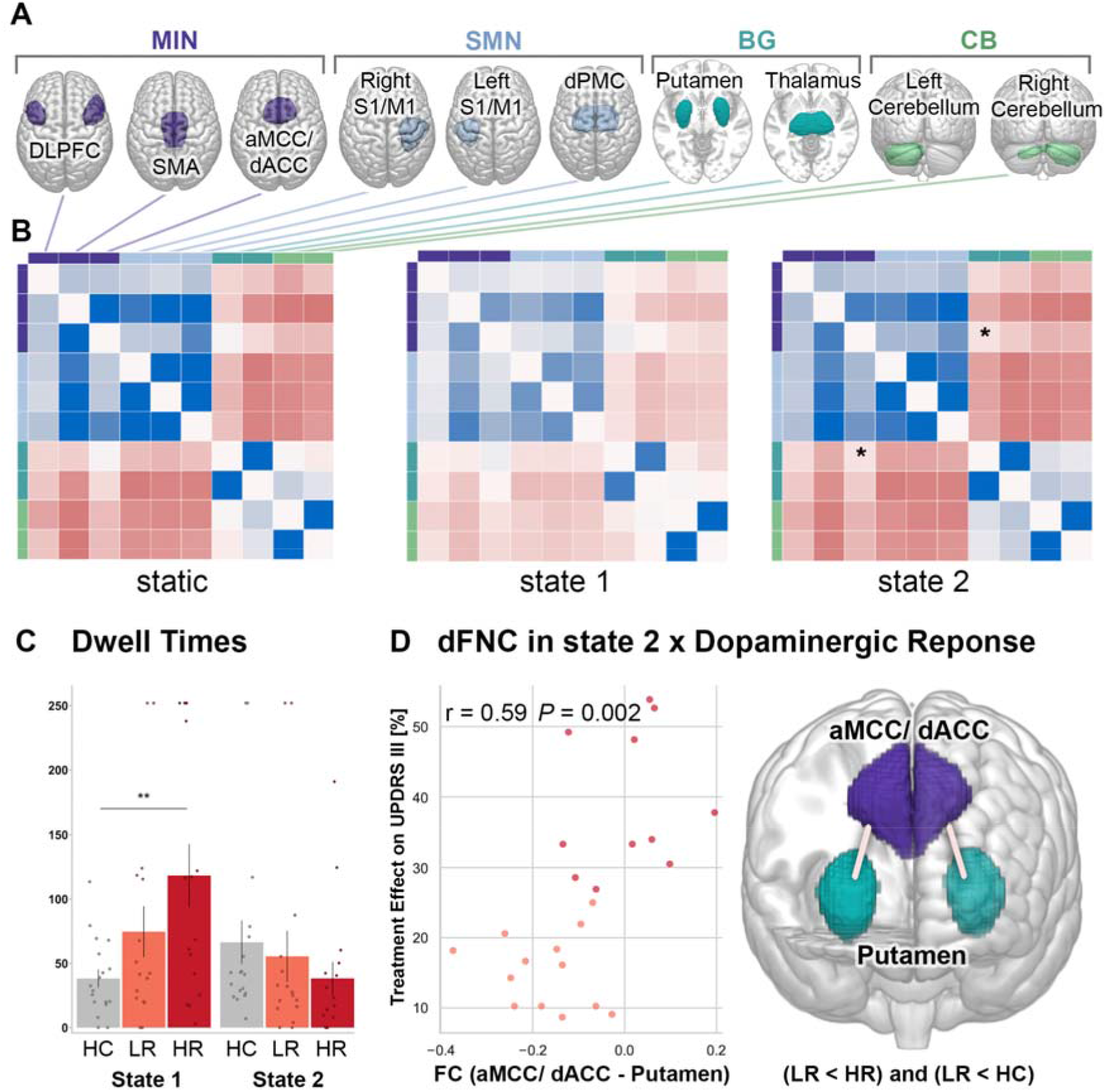
**(A)** Spatial ICA components across patients and healthy controls, allocated to color-coded networks: MIN (dark purple) = motor initiation network, SMN (light blue) = sensorimotor network, BG (dark green) = basal ganglia, CB (light green) = cerebellum. **(B)** Connectivity matrices across all healthy controls and patients indicating low (red) and high (blue) connectivity. Asterisk indicates significant group differences (FDR-corrected ANOVAs) **(C)** ANOVA results of dwell times showing that high responders spent more time than healthy controls in state 1 before changing to state 2. **(D)** The association between aMCC/dACC-putamen connectivity in state 2 and response to dopaminergic therapy is demonstrated by a positive pearson correlation (left) and significant group differences (right), indicating lower connectivity of low responders, than high responders and healthy controls. Abbreviations: HC = healthy controls, LR = low responders, HR = high responders. DLPFC = dorsolateral prefrontal cortex, SMA = supplementary motor area, aMCC/dACC = anterior midcingulate cortex/ dorsal anterior cingulate cortex, S1/M1 = primary sensorimotor cortex, dPMC = dorsal premotor cortex.

Static connectivity was assessed computing mean Pearson’s pairwise and Fisher z-transformed correlations. DFNC between the same VOIs was performed by applying the sliding window approach (13,20–22) in steps of one TR using a window size of 44 s and a Gaussian window alpha of three. As for static connectivity, all matrices resulting from the dFNC analysis were Fisher z-transformed. Matrices from all participants were finally entered into a k-means clustering analysis, grouping re-occurring connectivity patterns (13,21,23). The similarity of each window’s connectivity to the cluster centroid was estimated using the l1 distance (Manhattan distance). An optimal clustering solution of k = 2 was indicated by the silhouette measure (24) and elbow criterion (25).

We investigated the data using python (Version 3.8) including the ‘SciPy’ package (26). To measure the dynamic shifts between connectivity states, we assessed dwell times (mean time in one state without transitioning to another), fraction times (portion of total time in one state), and the number of transitions between states. Differences between these measures were compared by three-level one-way analyses of variance (ANOVAs) testing for group differences between the three groups: healthy controls, PD patients with low and high response to dopaminergic therapy. Likewise, we tested for group differences in connectivity pairs (static and dynamic), using three-level one-way ANOVAs for each connection (significance level P = 0.05, FDR-corrected).

## Results

Demographic and clinical information including testing for group differences is summarized in Table 1. Subgroups with low or high response to dopaminergic therapy did not differ in age, gender, motor or cognitive impairment, LEDD or within-scanner head motion.

**Table 1:** Demographics, clinical and MRI characteristics. Abbreviations: GDS-15 = Geriatric Depression Scale, HC = healthy controls, HR = high response to dopaminergic therapy, LEDD = levodopa daily equivalent dose, LR = low response to dopaminergic therapy, MDS-UPDRS III = Movement Disorder Society Unified Parkinson’s Disease Rating Scale part III, MoCA = Montreal Cognitive Assessment, NPT = Neuropsychological testing, PD = Parkinson’s disease, * = ordinal data is described by medians and interquartile range between first and third quartiles, instead of means and standard deviations. ^1^ANOVA, ^2^H-test, ^3^t-test, ^4^U-test.

|  | HC<br>(n = 18) | PD LR<br>(n = 16) | PD HR<br>(n = 16) | P |
| --- | --- | --- | --- | --- |
| Age [y] | 67.6 ± 7.7 | 66.6 ± 8.4 | 65.8 ± 7.0 | 0.784<sup>1</sup> |
| Sex [m/f] | 13/5 | 10/6 | 14/2 |  |
| Hoehn & Yahr stage* |  | 2.00<br>(2.00 - 2.25) | 2.00<br>(2.00 - 2.63) |  |
| Motor symptom<br>duration [y] |  | 4.9 ± 3.2 | 6.0 ± 3.3 |  |
| MDS-UPDRS III (ON)* |  | 29.00<br>(18.00 - 36.25) | 26.00<br>(18.00 - 28.25) |  |
| MDS-UPDRS III<br>(OFF)* |  | 35.00<br>(21.75 - 42.25) | 42.50<br>(27.75 - 51.00) |  |
| LEDD |  | 442.5 ± 259.8 | 659.6 ± 480.0 |  |
| GDS-15* | 0.50<br>(0.00 - 1.00) | 1.00<br>(0.00 - 2.00) | 0.00<br>(0.00 - 1.25) |  |
| MoCA* | 27.00<br>(26.00 - 27.00) | 26.50<br>(25.75 - 29.00) | 27.50<br>(25.00 - 28.00) |  |
| NPT [z-score] | 0.17 ± 0.47 | 0.33 ± 0.43 | 0.25 ± 0.46 | 0.628<sup>1</sup> |
| Rotation [degrees] | 0.0022 ± 0.0013 | 0.0021 ± 0.0007 | 0.0031 ±<br>0.0022 | 0.150<sup>1</sup> |
| Translation [mm] | 0.26 ± 0.08 | 0.25 ± 0.12 | 0.29 ± 0.10 | 0.480<sup>1</sup> |

Static functional connectivity across all participants showed positive connectivity within cortical, basal ganglia and cerebellar networks, but negative connectivity between cortical and subcortical networks (Figure 1 B). This connectivity pattern is in line with previous rs-fMRI studies on PD (8,10,27). FDR-corrected ANOVAs for *static* connectivity pairs did not show significant group differences. With regards to dFNC, we found two states, which essentially differed by their within- and between-network connectivity. Both states (Figure 1 B) featured positive within-network connectivity and negative between-network connectivity in cortical and subcortical networks. Yet, state 1 expressed this modular organization less pronounced, showing most positive connections within cortical, basal ganglia and cerebellar networks. In contrast, state 2 was marked by highly negative connections, segregating cortical regions from subcortical as well as cerebellar regions. Comparing dwell times between groups revealed that patients with high response to dopaminergic therapy spent significantly more time in state 1, compared to healthy controls (three-level one-way ANOVA: *P =* 0.010, *post-hoc* independent t-tests: *t*(32) = 3.3, *P* = 0.002, Figure 1 C). No significant differences were found for fraction times (*P =* 0.090) and number of transitions (*P =* 0.216). Furthermore, FDR-corrected ANOVAs indicated that dFNC between aMCC/dACC (anterior midcingulate cortex/dorsal anterior cingulate cortex) and putamen was significantly lower in patients with low response to dopaminergic therapy during state 2 and correlated with the effect of dopaminergic treatment on motor performance (Figure 1 D).

## Discussion

The present study revealed links between dynamic connectivity patterns and patients’ individual responses to dopaminergic therapy, which could not be detected by a comparable static connectivity analysis. Specifically, patients with high response to dopaminergic therapy spent more time in a state featuring lower within-network connectivity, but higher connectivity between networks (i.e., between cortical and striatal regions) - in other words, a more integrated brain state. The predominance of this state in high responders indicates that engagement of non-striatal and non-dopaminergic networks may foster a greater treatment response.

This finding can be interpreted in the framework of large-scale functional network organization (28–30), balancing between functional segregation and integration to enable optimal information processing. In this sense, high responders show a prolonged processing of information in a more integrative connectivity state, marked by less isolated connectivity within domains (Figure 1 C). Previous dFNC studies in PD associated a similar shift towards states with higher cortico-striatal integration with better cognitive (8,10) and motor (10) function, showing an association between such a dFNC fingerprint and a more benign PD phenotype.

Similar relationships between better clinical performance and network integration have been observed in other disease entities including stroke (31) and traumatic brain injury (32). These observations have led to the hypothesis that disturbed brain networks promote a more isolated within-network processing, which may be less prone to erroneous interactions with other domains (31). Particularly for PD, this may relate to animal models findings showing that dopamine depletion leads to a ‘loss of segregation’ in PD (33). Our findings suggest that a more integrated (thereby less segregated) connectivity between networks in PD contributes to an enhanced propagation of dopaminergic treatment effects across brain networks. In line with this interpretation, patients with *low* response to dopaminergic therapy expressed a more negative connectivity between mid- and anterior cingulate cortex and putamen in the more segregated state 2, compared to both healthy controls and high responders (Figure 1 D). This connection is known to be a crucial part of dopaminergic cortico-striatal loops involved in motivation and motor initiation (5,6,34), and was previously found to feature reduced functional connectivity in PD (27). Assessing static functional connectivity in a sample of 19 patients with advanced PD (disease duration 4-22y), Akram and colleagues have reported a similar correlation between the improvement of motor symptoms and fronto-striatal connectivity (6). While static connectivity was not able to detect such a relationship in our sample, our findings suggest that a transiently reduced cortico-striatal integration involving the mid- and anterior cingulate cortex critically relates to the motor improvement induced by dopamine replacement therapy.

As a limitation, it should be taken into consideration that all data were acquired in the ON state. Consequently, we cannot distinguish whether 1) the negative connectivity between aMCC/dACC and putamen in low responders reflects an insufficient dopaminergic up-regulation of cortico-striatal loops correlating with the clinical response across patients, or 2) reflects a functional network alteration with lacking cortico-striatal integration leading to the low dopaminergic effect. We did not perform a standardized levodopa challenge to measure treatment response. However, we expect the ON state to give a realistic estimate of the patient’s individual treatment effect. Still, we cannot rule out a potential influence of different treatment regimens.

Our data suggest that alterations in temporal dynamics of fronto-striatal connectivity might represent a biomarker for treatment response in PD. Longitudinal studies are warranted that investigate the effect of disease progression on this parameter.

## Data Availability

All data produced in the present study are available upon reasonable request to the authors.

## Funding

LH, CG and GRF are funded by the Deutsche Forschungsgemeinschaft (DFG, German Research Foundation) – Project-ID 431549029 – SFB 1451 (project C05).

MS is funded by the Koeln Fortune Program / Faculty of Medicine, University of Cologne (grant number 453/2018), and the Else Kro□ner-Fresenius-Stiftung (grant number 2019_EKES.02)

CEJD is supported by the Clinician Scientist Program (CCSP) / Faculty of Medicine / University of Cologne, funded by the Deutsche Forschungsgemeinschaft (DFG, German Research Foundation, FI 773/15-1)

## Competing interests

The authors report no competing interests.

## Supplement

### Neuropsychological assessment

Cognition was assessed as recommended in the MDS diagnostic criteria for mild cognitive impairment in Parkinson’s disease (1). Tests included Verbal Learning and Memory Test and Rey-Osterrieth Complex Figure Test for memory domain; Trail Making test, digit span, and Stroop color and word test for working memory; phonemic verbal fluency (-s version), semantic verbal fluency (animals), and Trail Making Test for executive functioning; similarities from the German version of the Wechsler Adult Intelligence Scale, and Aphasie-Check-List for language abilities; Benton judgement of line orientation, and Rey-Osterrieth Complex Figure Test for visuospatial function. Z-scores for each cognitive domain were calculated based on published age corrected norms, if available, and from an in-house cohort of healthy subjects matched for age. A composite score of the average of the five domain z-scores was calculated to estimate global cognitive performance.

### Resting-State fMRI Preprocessing

Rs-fMRI data were preprocessed using Statistical Parametric Mapping (SPM12; Wellcome Centre for Human Neuroimaging, London, UK) implemented in Matlab version 2019b (Mathworks Inc.; MA, USA). Distortion correction was performed using FSL (FMRIB Software Library v6.0; https://fsl.fmrib.ox.ac.uk/fsl/fslwiki/FSL). The first five EPI volumes were discarded avoiding noise from magnetic field saturation. The remaining 295 volumes were corrected for head movements by spatial realignment to the mean image, normalized into MNI space using the unified segmentation approach (2), and smoothed with a Gaussian filter of 8 mm full-width-at-half-maximum.

## Notes

### Competing Interest Statement

The authors have declared no competing interest.

### Author Declarations

The ethics committee/IRB of the University Hospital Cologne gave ethical approval for this work.

